# Trends in patients’ willingness for cancer care and the number of registered cancer cases in Ehime Prefecture during the COVID-19 pandemic

**DOI:** 10.1101/2022.11.30.22282924

**Authors:** Norihiro Teramoto, Natsumi Yamashita, Yutsuko Ohira

## Abstract

**Background:** The COVID-19 pandemic has reduced diagnosed cancer cases worldwide. This study aimed to elucidate the recovery of cancer care from the COVID-19 pandemic in Ehime Prefecture, Japan.

**Methods:** This study collected data from the hospital-based cancer registry (HBCR) as well as the number of outpatients, medical information provision fee payments (MIP2), and second opinion patients (SOP) from the Council of Ehime Cancer Care Hospitals (ECCH). Then cancer care and patient requests for hospital transfers before and during the COVID-19 pandemic were analyzed.

**Result:** The HBCR from the ECCH comprises >80% of cancer cases in Ehime Prefecture. In 2020, the numbers of all registered cases, first-line treatment cases, and cases detected by cancer screening in the HBCR decreased from those in 2018–2019. In 2021, they increased to almost the same levels as those in 2020. In contrast, the number of registered patients that changed hospitals (hospital-change cases) after first-line treatments, patients who lived outside the metropolitan area of Ehime but registered in metropolitan hospitals, MIP2, and SOP remained low in 2021 after decreasing in 2020. Furthermore, using the Wilcoxon rank sum test, the monthly numbers of hospital-change cases, MIP2, and SOP were significantly smaller in 2021 than in 2018–2019.

**Conclusion:** The assessed indicators suggest that the willingness of cancer patients to improve and/or advance cancer care had not returned to pre-pandemic levels by 2021. Hence, psychological measures in society and support for patient caregivers are necessary to prevent self-restraint in patients receiving cancer care.

**Mini-abstract:** The number of registered cases in hospital-based cancer registries returned to pre-COVID-19 levels by 2021, cancer patients’ willingness to further their care did not return to pre-pandemic levels.

## Introduction

Ehime Prefecture is one of the 47 local government areas in Japan, with a population of 1.34 million in 2019 and a nominal gross domestic profit (GDP) of 5 trillion yen, about one-hundredth of Japan’s population and GDP, respectively (1). Ehime Prefecture has problems common to the rest of Japan, such as a declining birth rate, an aging population, and a population concentration in the prefecture’s center. Approximately half of the residents live in the Matsuyama metropolitan area (Metro), which has a comprehensive cancer center, a university hospital, and the largest and second-largest general hospitals in the prefecture (2).

The Council of Ehime cancer care hospitals (ECCH) comprises 7 hospitals designated by the Ministry of Health, Labour and Welfare of Japan (MHLW) and 8 designated by the Ehime prefecture (3, 4). Although apart from Shikoku Cancer Center, these hospitals do not specialize in cancer care, they are recognized by the MHLW and/or Ehime Prefecture to meet the standards for staff and facility quality and the number of cancer treatments. Therefore, the ECCH-designated hospitals are expected to be the main institutes for cancer treatment in their medical districts. ECCH hospitals must have a hospital-based cancer registry (HBCR) (5).

The ECCH annually publishes “*Cancer Care in Ehime Prefecture Visualized by Cancer Registry* “ in print and on a website (2) to evaluate the status of cancer care in Ehime Prefecture using the Ehime prefectural hospital-based cancer registries (EHBCR). EHBCR accumulated since 2012. It is usually the earliest publication on the prefectural cancer registry results of the preceding year in Japan. In the recent 2 years, we also released *“COVID Bulletin 2020: Cancer Care in Ehime Prefecture Visualized by Cancer Registry”* and “COVID Bulletin 2021” as the earliest summary of HBCR in Japan to describe the cancer care during the pandemic (6, 7).

This publication shows the centralization of cancer care to the hospitals in Metro for some cancers, such as lung, breast, and gynecologic cancers (2).

Most patients’ cancer treatment in Ehime prefecture is completed at the ECCH hospitals, with few patient requests for transfers to hospitals in other prefectures (4). According to the population-based cancer registry of Ehime, a part of Japan’s National Cancer Registry, 81.2% of all cancers diagnosed in Ehime prefecture in 2017–2018 were diagnosed and/or treated in some ECCH hospitals (8). Furthermore, active first-line cancer treatment with surgery and/or chemotherapy is infrequent in non-ECCH hospitals. The COVID-19 pandemic is known to have reduced the number of cancer cases diagnosed and treated worldwide (9). The COVID-19 pandemic also impacted cancer care in Japan in 2020 because important medical procedures such as cancer screening and endoscopic examinations were restricted (10, 11). However, the restriction of medical treatment is not solely due to a lack of medical resources; there is also the pandemic’s influence on the social psychology of cancer patients (12-14). Hence, the restriction of medical treatment due to medical resource problems and the pandemic’s influence on cancer patients’ social psychology needs to be elucidated, although in many respects, cancer care in Japan rebounded in 2021 (2, 6). We hypothesized that the EHBCR and some additional data could elucidate cancer treatment in Japan as a one-hundredth microcosm.

## Material and Method

### Ehime prefectural hospital-based cancer registries

This study included the numbers of (i) all registered cases, (ii) patients receiving first-line treatment (first-line treatment cases), (iii) patients on watchful waiting as the first line of treatment (watchful-waiting cases), (iv) the cases detected by cancer screening (screening cases), and (v) patients who requested hospital transfers for further treatment after first-line treatments (hospital-change cases) in 2018–2021 from the EHBCR. The exclusion criteria included registered cases in the published EHBCR that were neither diagnosed, treated for cancer, nor followed up by watchful waiting in the hospitals. In addition, the cases were divided based on address into either Metro hospital cases of patients whose addresses were in Metro (M-M) or Metro hospital cases of patients who visited from the peripheral areas of Ehime or other prefectures (M-P). The other EHBCR data unshown in this report can be found on the ECCH website (2, 7).

### Other additional information from ECCH hospitals collected for the analysis

The monthly numbers of (i) outpatients from all the ECCH hospitals in 2018–2021 were collected, (ii) patients who were charged medical information provision fee II (MIP2), and (iii) patients who received a second opinion at the hospitals (second-opinion patients) (13, 15). MIP2 is a fee that hospitals charge patients needing a medical referral letter when requesting a second opinion in another hospital (MHLW, Table of Medical fees) (16). The number of MIP2 and second-opinion patients reflects the number of patients willing to visit medical doctors in other hospitals, hoping to receive better counseling for cancer care.

### Statistical analysis

The monthly numbers of the collected items were analyzed using the Wilcoxon rank sum test to compare the changes in cancer care in Ehime Prefecture before and during the pandemic. Differences were considered statistically significant at p-values <0.05. All statistical analyses were performed using Stata Version 17.0 (Stata Corp, College Station, TX, USA).

## Results

### Changes in the number of hospital-based cancer registrations in Ehime Prefecture before and during the COVID-19 pandemic

The numbers of all registered, first-line treatment, and screening cases in the EHBCR decreased in 2020 and increased in 2021 to almost the pre-COVID-19 levels (Table 1). The number of watchful-waiting cases increased in 2020 but decreased in 2021. Furthermore, the number of hospital-change cases decreased in 2020 and did not increase to pre-COVID-19 levels.

**TABLE 1.**
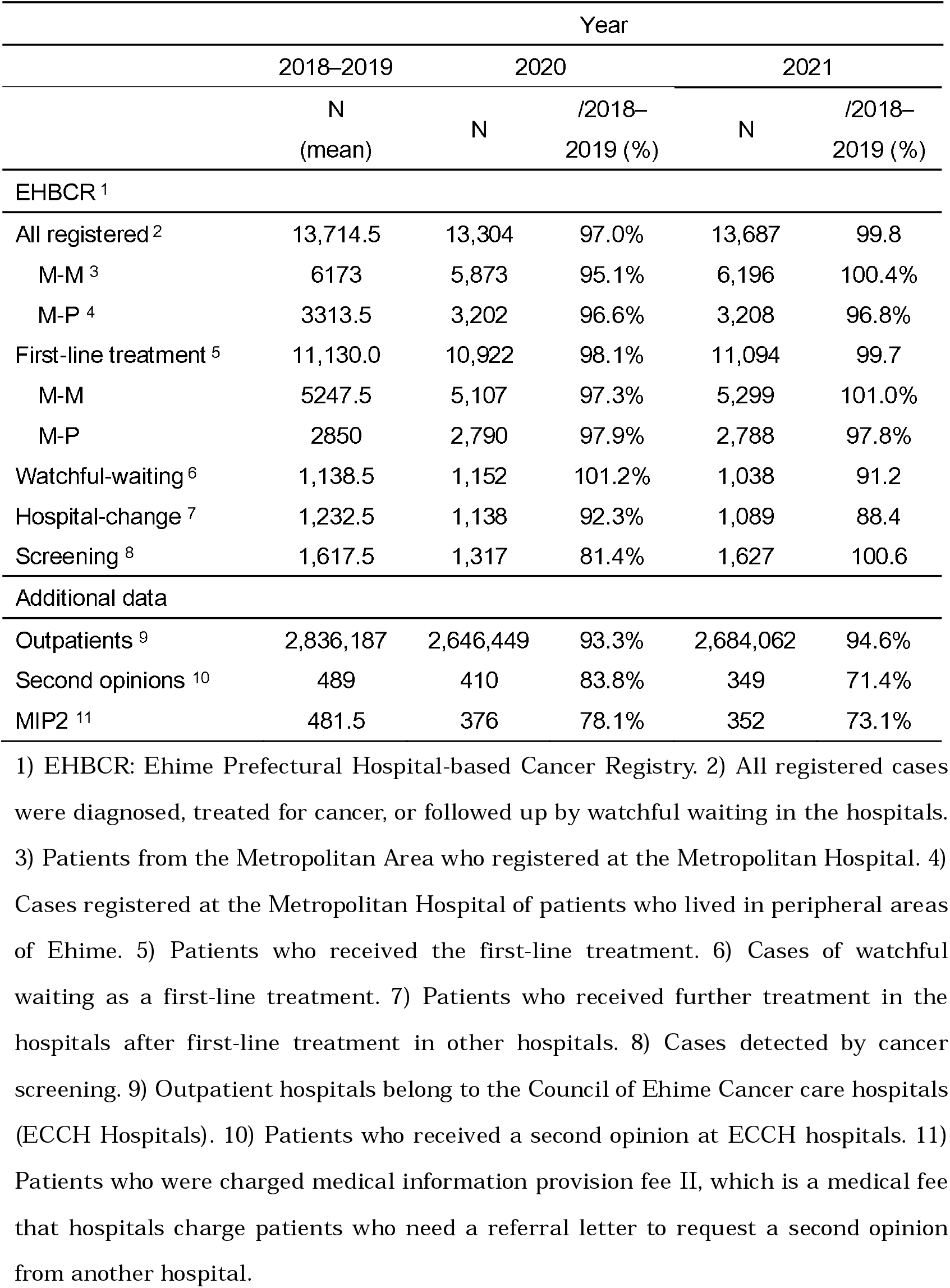
Annual summary of cancer care indicators in Ehime Prefecture before (2018–2019) and during the COVID-19 pandemic (2020 and 2021)

All registered and first-line treatment M-M cases decreased to 95.1% and 97.3% in 2020 from 2018–2019 (mean); they recovered to 100.4% and 101.0%, respectively, in 2021. In contrast, those of M-P cases decreased to 96.6% and 97.9%, respectively, in 2020. Unlike the M-M cases, no recovery was observed in 2021 (96.8% and 97.8 %, respectively) (Table 1).

The monthly numbers of all registered, first-line treatment, and screening cases decreased largely in May 2020 (Figure 1), when the Japanese government declared the first state of emergency. However, all registered and first-line treatment cases immediately increased to the normal annual levels, and the number of screened cases recovered relatively slowly until the end of 2020 (Figure 1). In December 2020 and January 2021, when the number of people infected with COVID-19 increased in Japan, the number of registered, first-treatment, and screening cases decreased again. However, they rapidly returned to the normal pre-COVID-19 monthly numbers. Meanwhile, the monthly number of hospital-change cases decreased in 2020 and did not return to the pre-COVID-19 level.

**Figure 1.**
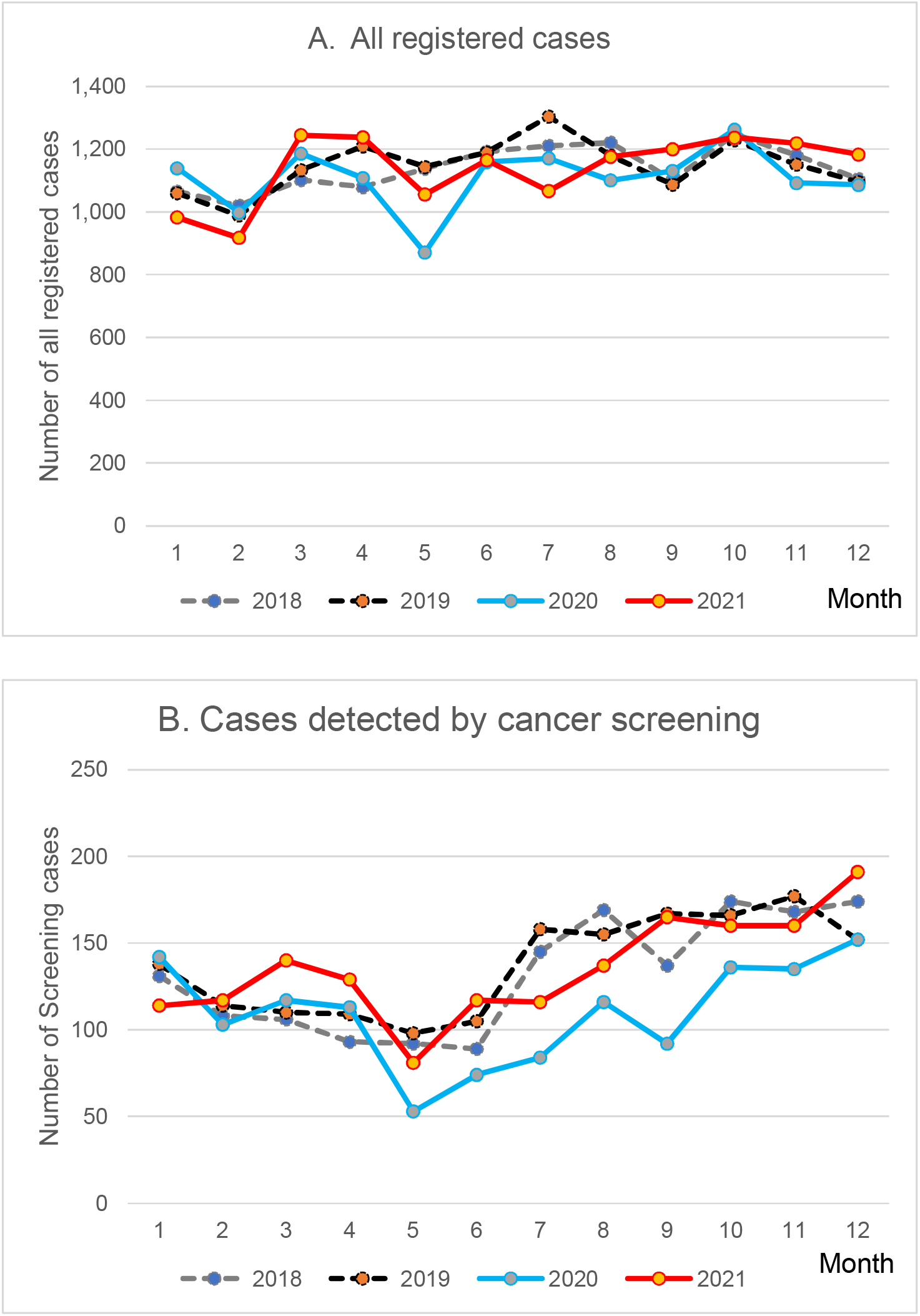
Monthly numbers of registered cases in Ehime before and during the COVID-19 pandemic The lines and dashed lines indicate the monthly numbers of registered cases each year. a) Line graph of all registered cases: there was a large drop in May 2020 (blue line), and the number returned to normal around September 2020. The red line (2021) runs with the dashed lines (2018–2019). b) Line graph of the number of cases detected by cancer screening: As with all registered cases, there was a large reduction in May 2020. However, it took longer to return to the pre-COVID-19 levels by the end of the year than the number of all registered cases (blue line).

In the Wilcoxon rank sum test, the monthly number of screening cases was significantly smaller in 2020 (median: 114.5, p=0.044) than in 2018–2019 (137.5); however, it increased in 2021 (133, p=0.875) (Table 2). The changes in the monthly numbers of all registered and first-line treatment cases before and during the pandemic were insignificant (Table 2). In comparison, the numbers of hospital-change cases were relatively smaller in 2020 (median: 94 /month, p=0.071) and significantly smaller in 2021 (91.5, p=0.018) than in 2018–2019 (103) (Table 2).

**TABLE 2.**
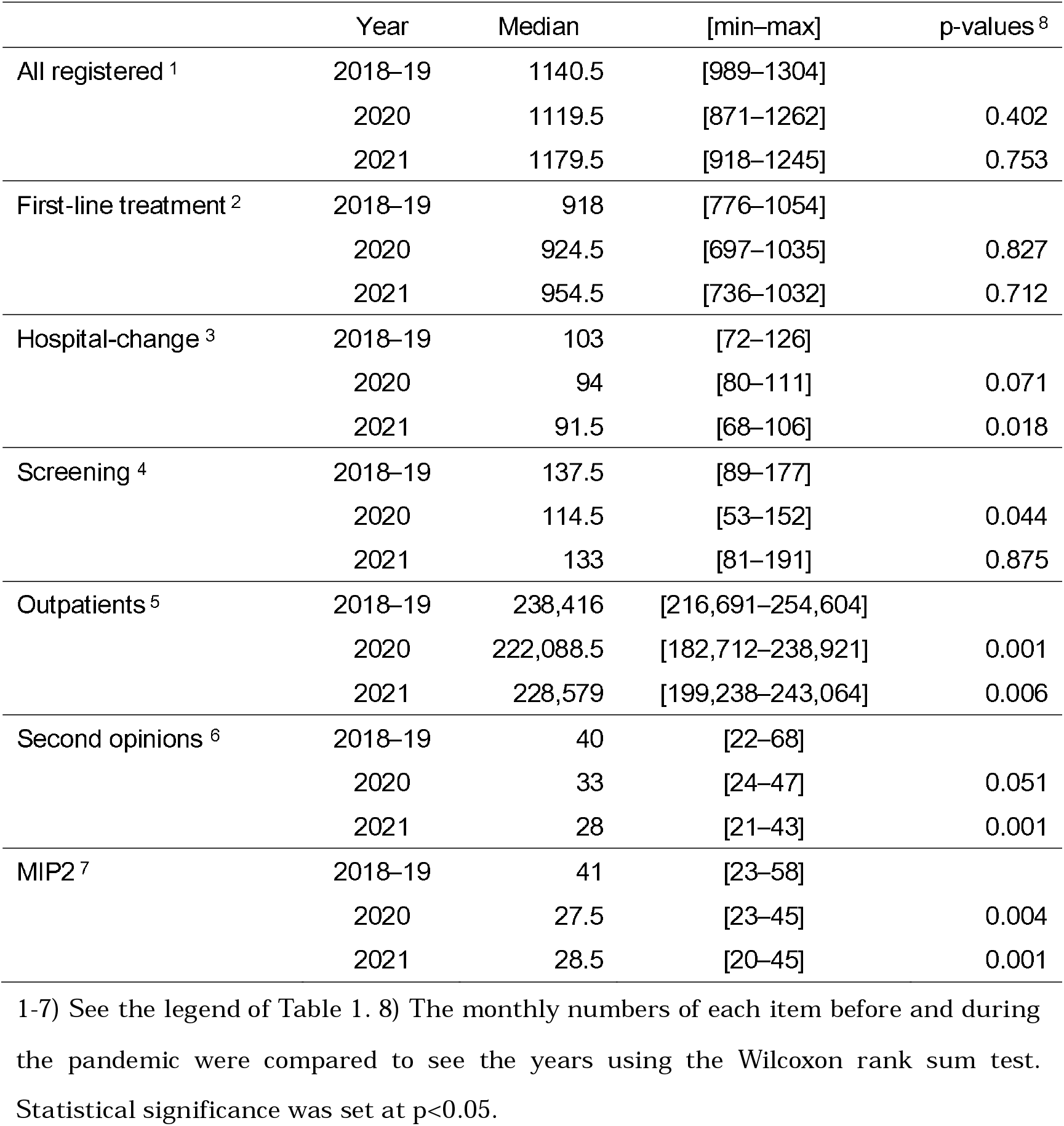
Monthly summary of cancer care indicators in Ehime Prefecture before (2018–19) and during the COVID-19 pandemic (2020 and 2021)

|  | Year | Median | [min–max] | p-values <sup>8</sup> |
| --- | --- | --- | --- | --- |
| All registered <sup>1</sup> | 2018–19 | 1140.5 | [989–1304] |  |
|  | 2020 | 1119.5 | [871–1262] | 0.402 |
|  | 2021 | 1179.5 | [918–1245] | 0.753 |
| First-line treatment <sup>2</sup> | 2018–19 | 918 | [776–1054] |  |
|  | 2020 | 924.5 | [697–1035] | 0.827 |
|  | 2021 | 954.5 | [736–1032] | 0.712 |
| Hospital-change <sup>3</sup> | 2018–19 | 103 | [72–126] |  |
|  | 2020 | 94 | [80–111] | 0.071 |
|  | 2021 | 91.5 | [68–106] | 0.018 |
| Screening <sup>4</sup> | 2018–19 | 137.5 | [89–177] |  |
|  | 2020 | 114.5 | [53–152] | 0.044 |
|  | 2021 | 133 | [81–191] | 0.875 |
| Outpatients <sup>5</sup> | 2018–19 | 238,416 | [216,691–254,604] |  |
|  | 2020 | 222,088.5 | [182,712–238,921] | 0.001 |
|  | 2021 | 228,579 | [199,238–243,064] | 0.006 |
| Second opinions <sup>6</sup> | 2018–19 | 40 | [22–68] |  |
|  | 2020 | 33 | [24–47] | 0.051 |
|  | 2021 | 28 | [21–43] | 0.001 |
| MIP2 <sup>7</sup> | 2018–19 | 41 | [23–58] |  |
|  | 2020 | 27.5 | [23–45] | 0.004 |
|  | 2021 | 28.5 | [20–45] | 0.001 |

### Changes in the numbers of outpatients charged MIP2 and second-opinion patients in ECCH before and during the COVID-19 pandemic

The numbers of charged MIP2 and second-opinion patients decreased in 2020 and further in 2021 (78.1% to 73.1% and 83.8% to 71.4%, respectively) compared to the mean in 2018–2019 (Table 1). Outpatients increased slightly in 2021 (2,684,062) after a decrease in 2020 (2,646,449); however, the number remained small at 94.6% of the number of outpatients in 2018–2019.

The monthly number of outpatients, MIP2 payments, and second-opinion patients decreased from April 2020, just before the first state of emergency was declared, and generally remained lower than the pre-COVID-19 levels, with some monthly increases and decreases (Figure 2). The Wilcoxon rank sum test showed that compared to the monthly numbers of outpatients, the monthly numbers of MIP2 payments were significantly lower in 2020 than those in 2018–2019 (p=0.001 and 0.004) and 2021 (p=0.006 and 0.001), respectively. The monthly numbers of second-opinion patients in 2021 were also significantly fewer than those in 2018–2019 (p=0.001) (Table 2).

**Figure 2.**
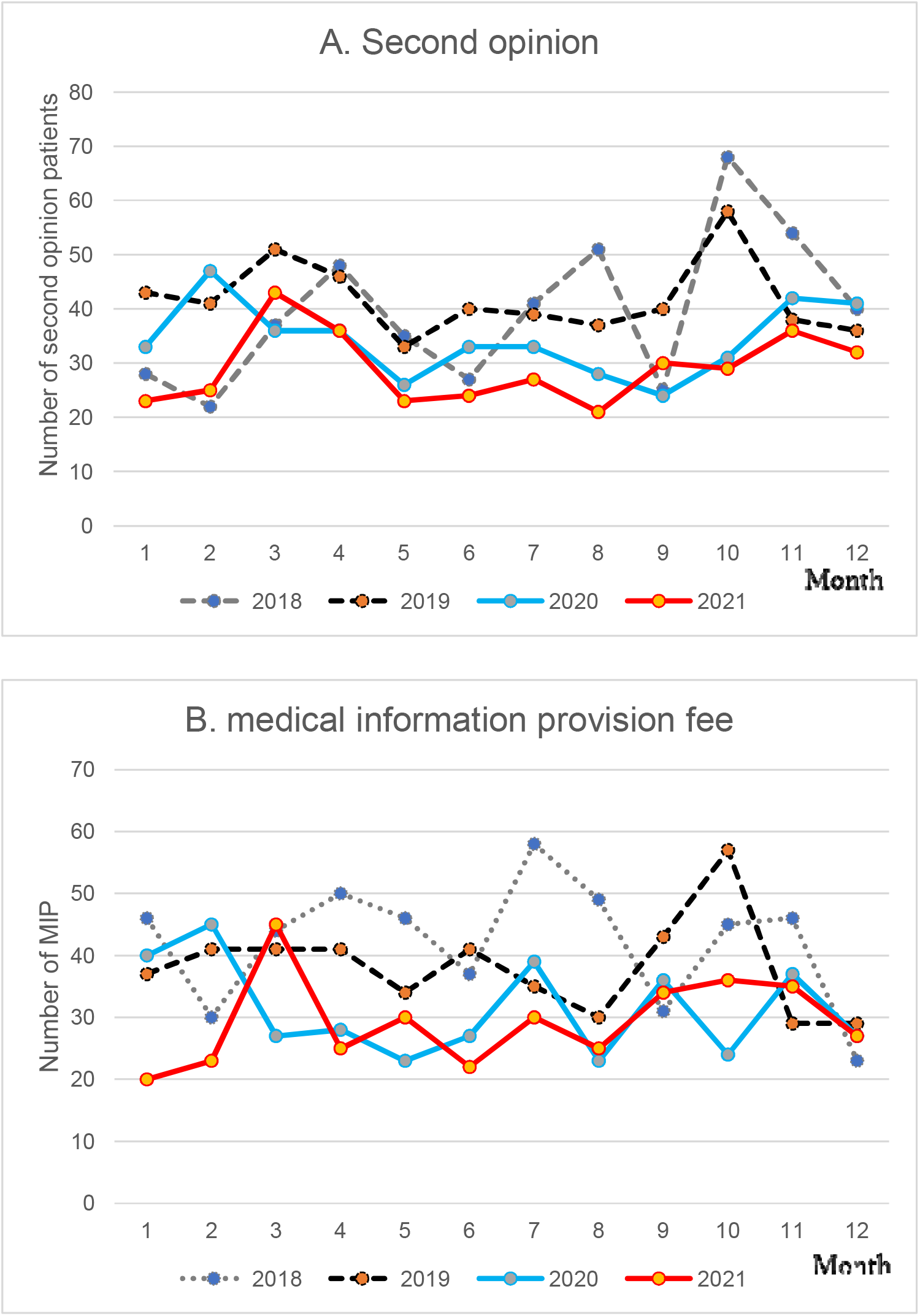
Monthly numbers of medical information provision II fee payments and second-opinion patients in Ehime cancer care hospitals before and during the COVID-19 pandemic Lines and dashed lines indicate the monthly numbers of second-opinion patients and medical information provision fee II (MIP2) payments each year. There was a large reduction in the number of second-opinion patients in May 2020 (a, blue line), and MIP2 payments decreased in April 2020 (b, blue line). Both run below the dashed lines (2018–2019) in most months through 2020 and 2021 (a, b).

## Discussion

In Ehime Prefecture, as well as in Japan, both the numbers of people infected with COVID-19 and related deaths increased in 2021 (17).

The numbers of registered, screening, first-line treatment, and active treatment (first-line treatment cases minus watchful-waiting cases) cases are indicators of cancer care activities in an area (5). The EHBCR of 2021 showed that these indicators increased from their 2020 levels to almost their pre-COVID-19 levels (Table 2). This suggests that primary cancer treatment in Ehime Prefecture recovered after the COVID-19 pandemic, presumably due to the establishment of cancer treatment systems, such as hospital-hospital collaboration and infection control measures at cancer screening institutions, during the pandemic.

On the other hand, the numbers of hospital-change cases, MIP2 payments, and second-opinion patients did not change in 2021, even after the recovery of the other indicators reflecting the number of active cancer treatments. Furthermore, although the numbers of all registered and first-line treatment of M-M cases recovered in 2021, those of M-P cases did not. This suggests that a proportion of cancer patients who live in the peripheral areas hesitated to undergo a trip to visit Metro hospitals, especially older and/or symptomatic patients (2).

One month after our report on Ehime Prefecture, the Center for Cancer registry of the National Cancer Center for Cancer Control of Japan released a rapid communication using National HBCR (NHBCR) (6). This report was in line with our results regarding the rebound in the numbers of registered, screening, and first-line treatment cases. In contrast, NHBCR did not show the change in numbers of hospital-change cases during the pandemic. It did not include the number of patients who visited hospitals in different medical areas. This might be due to the coverage ratio between the NHBCR and EHBCR. The rapid communication of the NHBCR used only the data from 455 hospitals which offered HBCR out of a total of approximately 850 hospitals. The remaining approximately 400 hospitals were excluded because they did not have the complete data sets from 2018-2021, or for other reasons. In addition, HBCR data of many smaller cancer care hospitals including 8 of 15 ECCH hospitals were not included in the NHBCR report. Therefore, EHBCR would better reflect local cancer care of Ehime prefecture.

The indices in this report were supposed to reflect the decline in patients’ willingness to receive cancer treatment (9, 13, 18). Although the pandemic was not severe enough to restrict patients’ hospital visits for primary cancer care in 2021, various psychological restraints, and restrictions on transportation for patients’ caregivers and visits by family and friends after hospitalization, might have caused a decline in patients’ desire to receive medical care. Hence, psychological measures and support for patient caregivers are necessary to prevent self-restraints due to the effects of the COVID-pandemic on the sentiments of patients with cancer toward receiving cancer care.

## Data Availability

This work was based on *COVID bulletin: Cancer Care in Ehime Prefecture Visualized by Cancer Registry*, one of the official operations of the Council of Ehime cancer care hospitals supported by Ehime prefectural government. The data included in this study are publicly available on the home page. All the data of EHBCR are available through Ehime Cancer Information Database Project if the application is approved to be relevant by IRB of Shikoku Cancer Center.

## Acknowledgments

This work is based on the hime Prefecture using the Ehime prefectural hospital-based cancer registries summarized annually by dedicated members of cancer registry subcommittee of Ehime Council of Cancer Care Hospitals (ECCH), including Junko Ohnishi (health information manager, HIM), Shizuka Kudara (HIM), Aoi Niida (HIM), Yoshiki Shiraoka (HIM), and Makoto Hamada (MD), Shikoku Cancer Center; Shoichi Matsukage (MD) and Hidekazu Shinjo (HIM), Uwajima City Hospital; Miyuki Yokoi, (tumor registrar, TR) and Mami Katakami (TR), Sumitomo Bessi Hospital; Miho Matsugi (HIM) and Takashi Matsumoto (MD), Ehime University Hospital; Nami Kusuhara (HIM), Takeshi Inoue (MD), and Masamitsu Tsubaki (HIM, MD), Ehime Prefectural Central Hospital; Tamura Junko (HIM) and Oobayashi Shohei (HIM), Japanese Red Cross Matsuyama Hospital; Sachiko Yano (HIM) and Syouta Tuduki (HIM), Saiseikai Imabari Hospital; Naomi Watanabe (HIM) and Tomomi Takahashi (HIM), Jyuzen General Hospital; Minako Kamada (TR) and Kayo Sasaki (HIM), Yawatahama City General Hospital; and the TRs of Hito Medical Center, Shikoku Central Hospital, Ehime Rosai Hospital, Saiseikai Saijo Hospital, Matsuyama Shimin Hospital, and Saiseikai Matsuyama Hospital. We also thank all members of the ECCH, especially its secretary Yoshie Takechi.

## DISCLOSURE

### ETHICAL APPROVAL

This research was approved by the institutional review board of Shikoku Cancer Center (Code: CO 2022-02), and was conducted in accordance with the Declaration of Helsinki.

### INFORMED CONSENT

Informed consent was not necessary in terms of the research ethics guidelines of Japan.

### REGISTRY AND THE REGISTRATION NO

N/A. This is not a clinical study.

### ANIMAL STUDIES

N/A. This is not an animal study.

## DATA AVAILABILITY STATEMENT

This work was based on “COVID Bulletin: Cancer Care in Ehime Prefecture Visualized by Cancer Registry,” one of the official publications of Ehime Cancer Care Hospitals (ECCH) supported by the Ehime Prefectural Government. The data included in this study are publicly available on the ECCH home page. All the data in the Ehime prefectural hospital-based cancer registries are available through *Ehime Cancer Information Database Project* if the application is considered to be relevant and is approved by the Shikoku Cancer Center IRB.

